# A study on control of novel corona-virus (2019- nCoV) disease process by using PID controller

**DOI:** 10.1101/2020.04.19.20071654

**Authors:** Habibolah Arasteh rad, Arshia Badi

**Affiliations:** Department of Complex Dynamics Modeling, Institute of applied Intelligent Systems, University of Tehran, Islamic Republic of Iran, Tehran

**Keywords:** Control Engineering, 2019-nCoV, SEIR Model, Quarantine, Social Distance

## Abstract

**Background:** In this paper, the SEIR dynamic model will be used to model the epidemic of coronvirus (2019-nCoV)disease. The SEIR model has been used to model infectious diseases in Malaysia.Then, the spread and control of the disease is simulated applying a PID controller. The results of this study show that the implementation of strict restrictions such as quarantine, social distancing and closure of gathering centers is effective in controlling the disease. Using the results and analyzing them, it was found that early and strict implementation of strict restrictions such as quarantine, social distance and closure of centers with a high percentage of community is very important to control this disease and prevent irreparable economic losses and depreciation of medical staff.

**Objective:** Modeling the prevalence and control of corona-virus (2019-nCoV)and the impact of government actions using control engineering methods.

**Method:** In this study, the SEIR dynamic model was used and the common data on the prevalence of the virus in Wuhan, China and Malaysia were used. As an example, the use of control target schemes is simulated in this paper.

**Results:** The findings of this study use control methods and forecasting in control engineering to provide a clear picture of macro-decisions for different governments in the field of infectious diseases.

**Conclusion:** Management and control schemes such as travel restrictions, quarantine, social distance and closure of offices, higher education institutions must be implemented immediately to prevent major economic and social losses. The implementation of these restrictions should not be delayed during the outbreak of corona-virus(2019-nCoV) infectious diseases.

## 1. Introduction

The end of 2019 and the beginning of 2020 saw the spread of a new virus. The World Health Organization has called the virus “2019-nCoV”[1]. One of the important effects of this virus was the impact on the lifestyle of human societies. On the other hand, there is a high epidemic of this virus, the control of which is complicated. Governments played a key role in countering coronavirus (2019-nCoV)in this campaign, from changing the health practices of communities such as emphasizing personal hygiene to very strict quarantine in Wuhan, China (the primary source of the disease) [2]. Disconnection between countries and closure of many sacred facilities in countries such as Iran [3], All of this was a sign of a new world of disease. According to the World Health Organization, due to the rapid transmission of the virus between humans, the only way to prevent the spread of coronavirus (2019-nCoV) is to cut off the human communication chain and maintain personal hygiene [1]. In this regard, countries have used various methods in this regard, for example, Iran has taken important steps to prevent the growth of the disease by closing many markets, higher education and implementing a social distance plan at the same time with the start of the New Year holiday [4]. South Korea is another country where people are exposed to the virus. Although the government constantly warned its citizens about health issues, there were no restrictions on travel in polluted areas, and even businesses continued to operate. In mid-February, the country considered the toughest quarantine measures for some cities and the northern province of Gyeongsang. Closing higher education centers and schools for long periods, preventing rallies and gatherings, using electronic maps to identify patients and knowing how they are moving, quarantining patients, allocating supplementary funding to fight corona virus (2019-nCoV), and raising alert levels Other Korean actions have been taken[7]. Hong Kong Despite its proximity to China and extensive ties to the country, it has also been successful in controlling the virus [5,6,7]. Hong Kong’s most important actions include;Strict regulations and heavy penalties for those who do not comply with the regulations. Allocating subsidies to all citizens to compensate for the closure of private businesses, Low-interest loans to large businesses and free distribution of masks and health goods to the poor population [7,8]. Another East Asian country has been affected by the Japanese virus, which, despite its vulnerable elderly population, has kept the virus-causing death rate very low. The Japanese with actions such as; Promoting telecommuting of large and small offices and companies, Assignment of some decisions to local governments in accordance with the instructions of the Ministry of Health, Long-term closure of schools and universities and subsidies for workers and employees to stay at home to care for children, Closing all gatherings and public places and increasing the capacity to perform diagnostic tests to about 4,000 cases per day, They were able to cope well with the spread of the virus [9,7].

In recent decades, mathematical models in epidemiology have been an important tool in the analysis of the spread and control of infectious diseases. The dynamic complexity of each disease dictates the use of simple mathematical models to gain insight into the spread of disease and test control strategies [10]. Since it was the first application of optimal control in biomedical engineering in the 1980s [11], several vaccination strategies for infectious diseases have been successfully modeled as optimal control problems. The so-called computational models have been widely used in this field. One of these models is SEIR, which is based on dividing the study population into four parts. The person may be susceptible (S) to the disease but not yet infected (E), infectious (I), or improved (immune) (R). SEIR models can show many infectious human diseases such as measles, smallpox, influenza, dengue, etc. [12], but in this article we will look at a general SEIR model. Generally, to achieve the goal in the shortest possible time, dynamic models use the principles of control engineering theory.

Control and feedback is one of the basic concepts of today’s world that is used in many areas. The word “control” has been used in many aspects of our lives. Financial control, community behavior, pest control, engine control, robot control, and hundreds of other examples are familiar with these applications. It can also be said “Power without control is worthless”. This statement is true in many areas, such as technology and social issues. For example, when a person feels cold (feeling the stage). It decides to wear and act on it (the decision-making stage and then the control action) and thus provides itself with warmer and more comfortable conditions (purpose). This is an example of biological feedback due to changes in the environment. [13,14]

This article includes five main sections, the first of which discusses the status of coronavirus (2019-nCoV) disease, the second section examines the modeling of infection in Malaysia as an example, and then introduces a classic engineering and applied controller. In the fourth section, we will implement and review the desired methods using computer simulation, and in the last section, the general results of this article will be examined.

## 2. Modeling

In this article, using the SEIR model of Malaysia[15], we will discuss the creation of a control solution using mathematical tools:

The transmission model is based on the epidemiological aspects of the disease [15].

- In this study, the Malaysian population is divided into SEIR models, which consist of four parts: susceptible (S), exposed (E), infected (I), and recovered (R). And the population is assumed to be unchanged. It is assumed that at first the whole population is susceptible, hence.
- Due to the growing number of cases around the world in a short period of time, the disease is assumed to be contagious enough.
- Birth and natural death rates have not been seen in this model.
- It is assumed that 2019-nCoV is capable of transmitting between humans.

Differential equations (equations (1) - (4)), which describe the dynamics of 2019-nCoV in the human population, are formulated based on a diagram first described in Figure (1). [15]

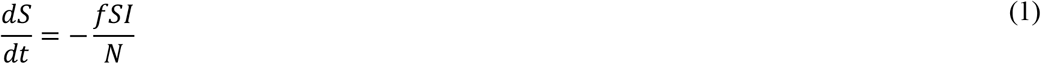

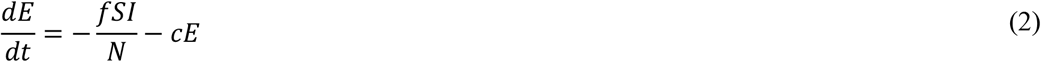

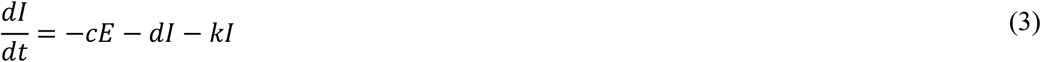

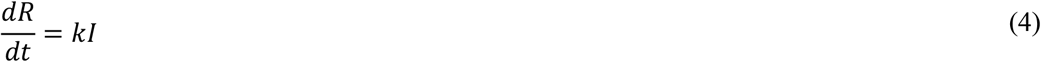

In the above figure model, since zero of the patient may be infectious and moving and freely interact with susceptible humans, Therefore, susceptible humans are exposed to this disease at the rate of transmission (f). After exposure, the latent period begins, and after this period, These infections and contaminants are then transferred to the infected area at a rate of (c) and when they recover from the disease, they are transferred at an improved rate (k).

Once exposed, the incubation period begins, and after this period, these motions and groups that are exposed to contamination are transferred to the infected part at a rate of (c) and when they recover from the disease, at a rate of (k).) Are transferred to the improved part. However, there are some contaminated items that may die due to disease with a certain rate (d)[15].

## 3. PID controller design

PID control is one of the most popular types of controllers used in industrial applications, and more than 90% of all controllers enforce PID or PI (unproductive) regulations [16,17]. Therefore, a desired state (ie, a set of points, reference, target) indicates the ultimate goal of the adjustment process.

PID controllers are based on closed-loop strategies with negative feedback mechanisms that track the actual state of the environment. In the most traditional implementation of negative feedback methods, the difference between the measured mode of the variable for adjustment (such as real temperature in a room) (e.g., real temperature in a room) and the target value (For example, 25° C) creates a prediction error that minimizes the controller output, e.g. If the temperature is too high, it will decrease and if it is too low, it will increase. Mathematically, we have the error as equation (4):

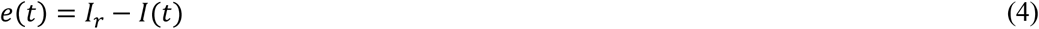

Where *e*(*t*)is an error, *y*_*r*_ is the reference or setting point (for example, the desired temperature) and *y*(*t*) is the observed variable (for example, the actual room temperature). This mechanism is unstable under very normal conditions, especially when a steady state compensation is added (such as sudden, unpredictable changes in external conditions that affect room temperature that we do not control) or when fluctuations are needed. Suppressed (such as over-oscillations, while temperature regulation may be undesirable). PID controllers Add shape 1 to the standard negative feedback architecture, here called the so-called P term, with an integral or I and derivative or term D, beautifully dealing with both problems, see Figure 1. Accumulates predictive error over time to deprive errors due to unstable input from computational mode, While minimizing predictive error derivatives leads to a reduction in the range of fluctuations of the controlled signal. The general shape of the *u*(*t*) control signal generated by a PID controller is usually determined by Equation (5).

**Figure 1:**
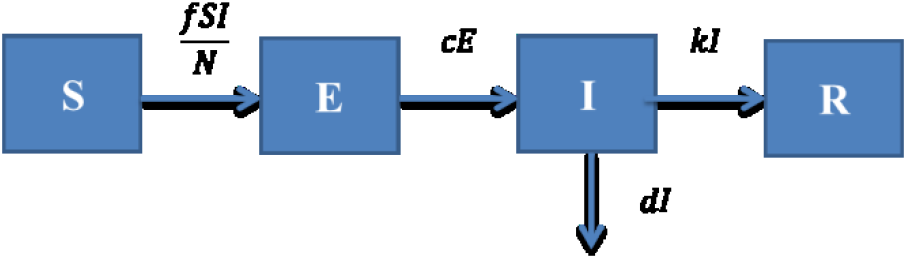
SEIR model based on reference [15]

**Figure 2:**
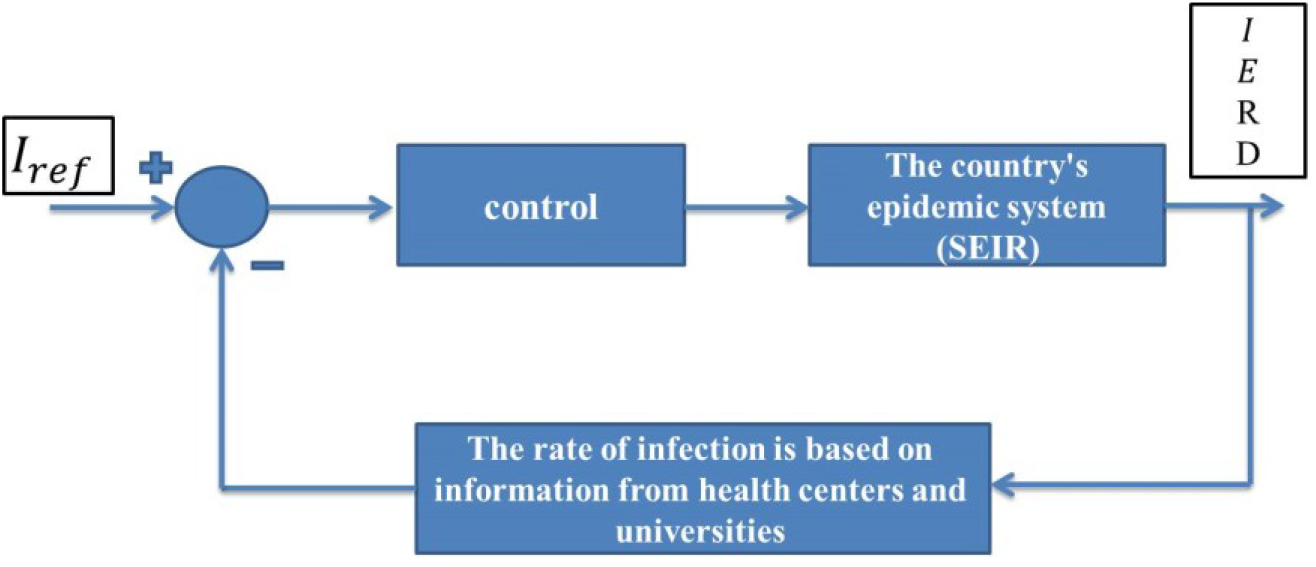
coronavirus (2019-nCoV) control system using PID controller

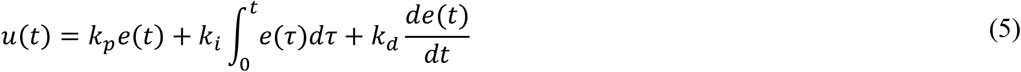

In the above equation e(t), the prediction error and *k_p_, k_i_, k_d_* are proportional, integral, and derivative, respectively, a set of parameters used to regulate the relative resistance of conditions P, I, and D. It can be said that the popularity of PID controllers is largely due to their simple formulation and implementation. Another important challenge is to adjust the parameters *k_p_, k_i_, k_d_*, which must be compatible with the various (often contradictory) limitations in the adjustment process [19,20].

## 4. Model simulation

To simulate the 2017 MATLAB software, we use the Simulink toolbox, the controller acts as a decision maker (government or health ministry in a country). The executive tools of this issue are the executive apparatus within a country, such as the Ministry of Health, medical centers, military forces, and relevant organizations. For example, quarantine is one of the most effective ways to reduce transmission and contact between people, and then vaccination is recommended to eradicate it. It should be noted that in real-world situations, costly decisions cannot be made because they may have economic and social consequences. This statement is like a control effort in a system, a controller with high control effort is practically useless.

The first confirmed Malaysian infectious disease began on January 23, 2020, when he returned from Singapore.Therefore, the initial time is January 23, 2020 with a positive case. In Tables (1, 2, and 3), we see the constant and initial parameters of the SEIR model, the controller parameters in the first scenario, and the controller parameters in the second scenario, respectively.

**Table 1:** SEIR model parameters

| Parameter | Description | Value | Source |
| --- | --- | --- | --- |
| $N$ | Total human population | 32600000 | [22] |
| $\frac{1}{c}$ | Incubation Period | 6.5 | [23] |
| $f$ | Transmission rate of I to S | 1.07 | [24] |
| $\frac{1}{k}$ | Infectious Period | 3.6 | [24] |
| $d$ | Death rate of 2019-nCoV | 0.03 | [15] |
| $E_0$ | Exposed | 0 | [15] |
| $I_0$ | Infected | 1 | [15] |
| $R_0$ | Recovered | 0 | [15] |

**Table 2:** Control parameters in the first scenario

| Parameter | Assumption | Value |
| --- | --- | --- |
| $k_p$ | Equivalent to early implementation of restrictive plans (closure of higher education, offices, market, disconnection between individuals) | 0.1 |
| $k_i$ | Effectiveness of quarantine or social distance, implementation of positive health plans | 0.1 |
| $k_d$ | Increase or decrease the time it takes to implement quarantine plans | 0.5 |

**Table 3:** Control parameters in the second scenario

| Parameter | Assumption | Value |
| --- | --- | --- |
| $k_p$ | Equivalent to early implementation of restrictive plans (closure of higher education, offices, market, disconnection between (individuals | **0.5** |
| $k_i$ | Effectiveness of quarantine or social distance, implementation of positive health plans | **0.1** |
| $k_d$ | Increase or decrease the time it takes to implement quarantine plans | **0.8** |

### Scenario (1): Achieving a goal with a fixed level

After 300 days of simulation, we see that in Figure (3, right) the prevalence of coronavirus (2019-nCoV), as shown in the reference, is estimated to be more than 3.5 million in the 90-to 100-day period. In this case, no control is observed. Using the proposed controller in Section (3), it is observed that the process of infection has been controlled with a positive sample from the beginning and the first wave of the disease, although it has severe shocks, can reduce the infection process with accurate and complete implementation of local quarantine schemes. To prevent the spread of the trend, this can be achieved with the help of government shutdowns, reduced traffic, higher education closures, and tighter urban traffic conditions. The highest incidence occurs in the first few days, which is due to the strict implementation of the mentioned control plans, we see a decrease in statistics. However, it should be considered that the stabilization of the rate means that the facilities, medical equipment and information related to the spread of the disease will be provided to the medical community of the country using the plan in a controlled manner (Figure (3, left)).

**Figure 3:**
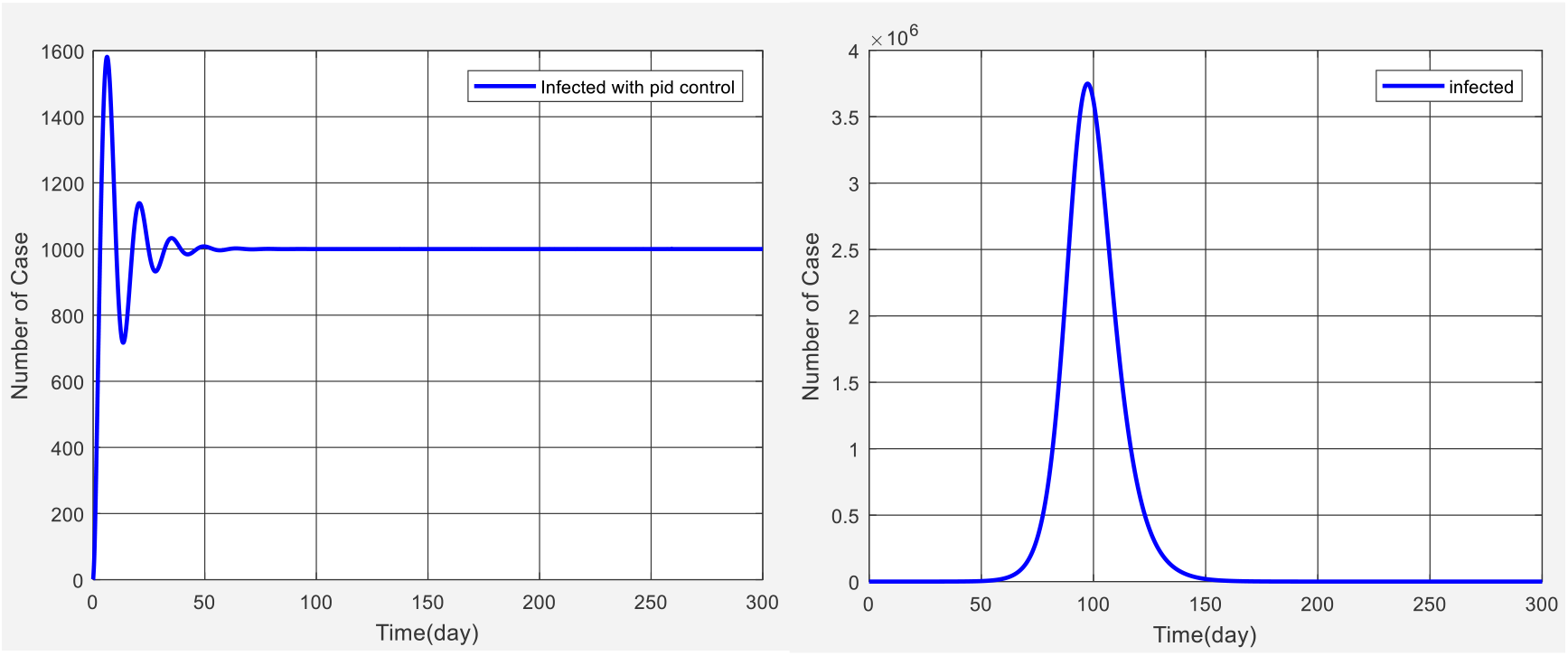
Right: SEIR modeling using reference [15], Left figure: Control system design to reduce patients and control them under a thousand people using PID controller

### Scenario (2): Variable goal based on time and government decision

In Figure (4, right) we see another example of a government in which, especially health centers, want to reduce the number of patients while increasing the number of patients to prevent further spread of the disease, Of course, it should be noted that this is a sample plan. In this plan, according to the process of the onset of the disease, it can be seen that the control system has been able to follow the goal well, which is specified in Figure (4, left). This important issue has put the disease under control in a predicted state, a plan similar to that proposed for Malaysia, but could include any other plan.

**Figure 4:**
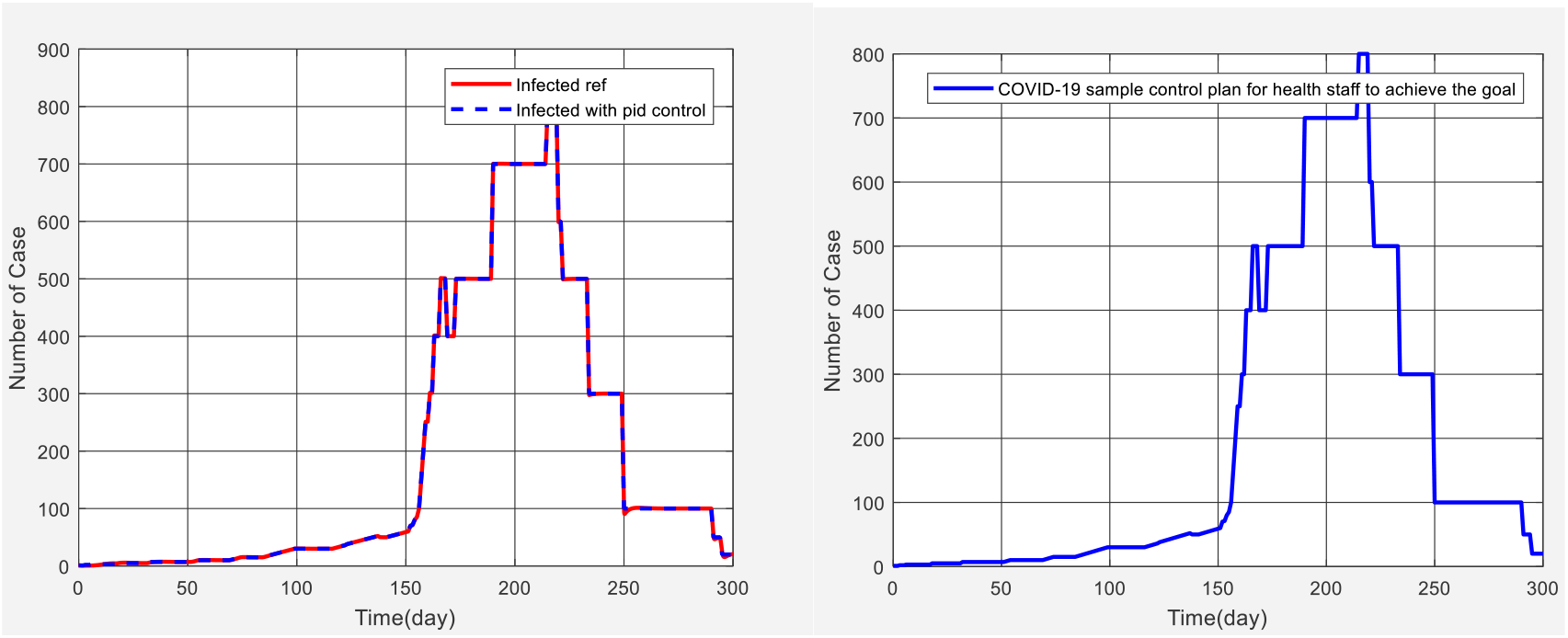
Right: Sample plan in which the government, especially health centers, wants to reduce the number of patients at the same time by increasing the number of patients to prevent further spread of the disease by considering the SEIR model,Left: Tracking the target using the PID controller

Figure (5, right) shows the control effort, which in the peak of the disease increases the performance of the controller and shows the unprecedented tightening in the country. Of course, the control effort in the field of engineering is very important because the excessive amount of rules and regulations cannot be done by the executive and operational agents with limited power, and the amount must be reasonably good as the country is dealing with good executive infrastructure. Also, the error in the control method is shown in Figure (5, left), which is only around the 250th day of this value due to the peak of the disease and shows the success of the controller in this matter.

**Figure 5:**
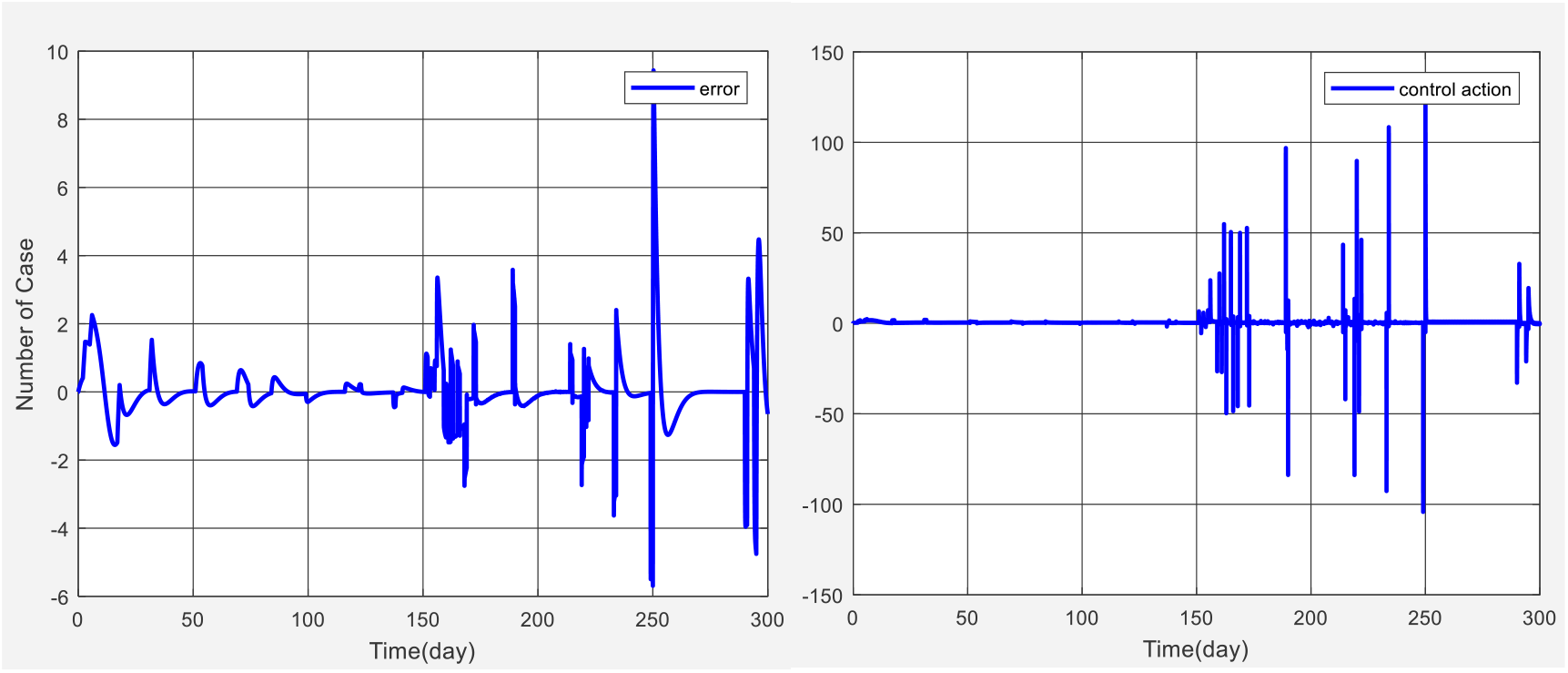
Right: Control effort, Left: Tracking error

## 5. Conclusion

In this paper, we have tried to design a suitable controller to prevent the spread of coronavirus (2019-nCoV) disease. The prevalence of Covid-19 virus has been modeled by SEIR, then it has been tried to apply government control decisions to the model with PID controller and measure the prevalence of the disease by changing the difficulty of controls. The results of the study show that if no control is performed, the rate of patients with 2019-nCoVwill spread to more than 35% of the population within 100 days of seeing the first patient. By using controllers, the infection rate has been controlled since the first positive sample was recorded and the infection rate has decreased. As the level of control strictures and restrictions increases, the rate of contagion of the disease has dropped dramatically.

## Data Availability

Information related to the study is in the manuscript

## Abbreviations

Covid-19 coronavirus; basic reproduction number; Malaysia

## Availability of data and materials

Information related to the study is in the manuscript

